# Vitamin D, chronic pain, and depression: linear and non-linear Mendelian randomization analyses

**DOI:** 10.1101/2023.04.12.23288467

**Authors:** Emily Bassett, Eva Gjekmarkaj, Amy M. Mason, Sizheng Steven Zhao, Stephen Burgess

**Affiliations:** MRC Biostatistics Unit, University of Cambridge, Cambridge, CB2 0SR, United Kingdom; Department of Public Health and Primary Care, University of Cambridge, Strangeways Research Laboratory, Cambridge, CB1 8RN, United Kingdom; British Heart Foundation Cardiovascular Epidemiology Unit, Department of Public Health and Primary Care, University of Cambridge, Cambridge, CB2 0BD, United Kingdom; Centre for Epidemiology Versus Arthritis, Division of Musculoskeletal and Dermatological Science, School of Biological Sciences, Faculty of Biological Medicine and Health, University of Manchester, Manchester Academic Health Science Centre, Manchester, UK

## Abstract

Vitamin D deficiency has been linked to various chronic pain conditions. However, randomized trials of vitamin D supplementation have had mixed results. In contrast, systematic reviews of randomized trials indicate a protective effect of vitamin D supplementation on depression. We undertake a Mendelian randomization investigation in UK Biobank, a study of UK residents aged 40-65 at recruitment. We perform linear and non-linear Mendelian randomization analyses for four outcomes: fibromyalgia, clinical fatigue, chronic widespread pain, and probable lifetime major depression. We use genetic variants from four gene regions with known links to vitamin D biology as instruments. In linear analyses, genetically-predicted levels of 25-hydroxyvitamin D [25(OH)D], a clinical marker of vitamin D status, were not associated with fibromyalgia (odds ratio [OR] per 10 nmol/L higher 25(OH)D 1.02, 95% confidence interval [CI] 0.93, 1.12), clinical fatigue (OR 0.99, 95% CI 0.94, 1.05), chronic widespread pain (OR 0.95, 95% CI 0.89, 1.02), or probable lifetime major depression (OR 0.97, 95% CI 0.93, 1.01). In non-linear analyses, an association was observed between genetically-predicted 25(OH)D levels and depression in the quintile of the population with the lowest 25(OH)D levels (OR 0.78, 95% CI 0.64, 0.94); associations were null in other strata. Our findings suggest that population-wide vitamin D supplementation will not substantially reduce pain or depression; however, targeted supplementation of deficient individuals may reduce risk of depression.

## INTRODUCTION

Vitamin D deficiency, defined as low levels of circulating 25-hydroxyvitamin D (25(OH)D)^1^, has been linked to recurrent and chronic pain conditions. Profound vitamin D deficiency is strongly linked to pain from osteomalacia, a bone weakening disorder^2^. Low vitamin D levels have also been suggested to play a role in the aetiology of chronic pain states through their influence on bone biology, autoimmune diseases, inflammation, or neuromuscular and other immunological influences on pain manifestation^3, 4^. Empirical evidence from observational studies indicates hypovitaminosis D to be highly prevalent in patients with chronic pain^5^, fibromyalgia^6^, and depression^7^.

However, evidence for a direct causal link between low 25(OH)D levels and chronic pain conditions is incomplete. Randomized controlled trials examining the impact of vitamin D supplements as a treatment for chronic pain conditions such as fibromyalgia are limited in size and quality^8, 9^. In contrast, systematic reviews of randomized trials for depression^10^, depressive symptoms^11, 12^, and negative emotions^13^ have indicated beneficial effects of vitamin D supplementation, although no evidence for an effect was observed in the large Vitamin D and Omega-3 Trial (VITAL)^14^. Inverse associations have also been observed in observational epidemiological studies^15-17^, although such investigations are vulnerable to bias from confounding and reverse causality: individuals with depression could have low vitamin D levels due to spending less time outdoors or changes in appetite and diet.

An alternative approach to randomized controlled trials that addresses the issues of confounding and reverse causality is Mendelian randomization. Genetic variants having a specific association with circulating 25(OH)D levels can be used as unconfounded proxies for changes in 25(OH)D levels to assess the likelihood of a causal link between vitamin D and chronic pain conditions. As genetic variants are randomly distributed at conception, they are unlikely to be related to possible confounders^18^. In addition, chronic pain conditions cannot affect the genotype and so reverse causality is avoided^19^. As a result, Mendelian randomization analyses can provide more reliable insights into any potential causal relationship between vitamin D and chronic pain conditions compared with conventional observational studies. As the shape of the relationship between 25(OH)D serum levels and chronic pain conditions is unknown, and many observational vitamin D studies have shown a threshold effect^5^ – where the disease risks are only related to the people with vitamin D deficiency – the standard Mendelian randomization approach which assumes linearity may overlook non-linear effects of 25(OH)D.

Given the uncertainty in the empirical literature as to the nature of the relationship between vitamin D serum levels and chronic pain conditions, we investigated evidence for a causal effect of 25(OH)D concentrations on chronic pain outcomes. We used individual-level data from the UK Biobank study to perform both linear and non-linear Mendelian randomization analyses. We also considered depression as an additional outcome, to validate trial findings and investigate the evidence for a causal effect on this outcome both in the population as a whole, and for strata of the population to assess the shape of any potential effect.

## SUBJECTS AND METHODS

### Study population

The UK Biobank cohort comprises around 500,000 participants aged 40 to 69 years at baseline, recruited between 2006-2010 at 22 assessment centres throughout the UK, and followed up for a variety of health conditions from their recruitment date until July 2019 or their date of death. Informed consent was obtained from all participants. The dataset includes genome-wide genotyping of baseline samples from all participants, results of clinical examinations, assays of biological samples, detailed information on self-reported health behaviour, and is supplemented by linkage with electronic health records such as hospital inpatient data, mortality data, and cancer registries^20^.

We restricted our attention to 333,025 unrelated participants of European ancestry that passed various quality control tests and had a valid 25(OH)D measurement. European ancestry was defined using self-reported ethnicity and genomic principal components as described previously^21^. We removed related individuals so that only one person in each family (defined as third-degree relatives or closer) was included in the analysis.

### Vitamin D measurement and classification

Concentrations of 25(OH)D in blood were measured using the DiaSorin Liaison immunoassay analyser. Measurements were mean-shifted to correspond to a measurement taken in October, in order to account for seasonal variability in 25(OH)D concentrations.

### Outcomes

We considered four outcomes: fibromyalgia, clinical fatigue, chronic widespread pain, and probable lifetime major depression.

Fibromyalgia was defined as ICD-9 code 729.1, ICD-10 code M79.7, or self-reported code (field 20002) 1542. Clinical fatigue was defined as ICD-9 code 780.7, ICD-10 code R53, or self-reported code 1482. Chronic widespread pain was defined as answering yes to the four questions “Have you had pain for more than 3 months?” relating to the neck or shoulder (field 3404), back (field 3571), hip (field 3414), and knees (field 3773).

Probable lifetime major depression was defined as the presence of four criteria^22^: 1) answering yes to the question “Looking back over your life, have you ever had a time when you were feeling depressed or down for at least a whole week?” or “Have you ever had a time when you were uninterested in things or unable to enjoy the things you used to for at least a whole week?”, 2) answering 2 weeks or more to the question “How many weeks was the longest period when you were feeling depressed or down?”, 3) answering 2 or more to the question “How many periods have you had when you were feeling depressed or down for at least a whole week?”, 4) answering yes to the question “Have you ever seen a general practitioner for nerves, anxiety, tension or depression?” or “Have you ever seen a psychiatrist for nerves, anxiety, tension or depression?”.

### Genetic variants

To minimize potential bias due to horizontal pleiotropy, we took genetic variants from four gene regions previously shown to be strongly associated with 25(OH)D and implicated in the transport, metabolism, and synthesis of vitamin D ^23^ – *GC, DHCR7, CYP2R1*, and *CYP24A1*. The *GC* gene encodes vitamin D binding protein. The *DHCR7* gene product converts 7-dehydrocholesterol to cholesterol, reducing 7-dehydrocholesterol available for conversion to previtamin D_3_ by solar radiation. The *CYP2R1* gene encodes vitamin D 25-hydroxylase, a regulator of 25(OH)D synthesis through 25-hydroxylation of vitamin D in the liver. The *CYP24A1* gene product inactivates the active form of vitamin D (1α25(OH)_2_D).

To maximize the variance explained by the genetic instrument, we considered available variants at each genetic locus, and selected variants associated with 25(OH)D concentrations using a stepwise selection method^24^. In total, 21 variants were included in the analysis (Supplementary Table 1). We constructed an allelic score weighting variants by their conditional associations with 25(OH)D concentration.

### Statistical analyses

#### Linear analyses

Linear Mendelian randomization estimates were calculated using the ratio method by dividing the genetic association with the outcome by the genetic association with 25(OH)D concentration, and scaling the estimate to a 10 nmol/L difference in genetically-predicted 25(OH)D concentration. Genetic associations were estimated using logistic regression for disease outcomes, and using linear regression for 25(OH)D concentrations. All regression models were adjusted for age at baseline, sex, centre, and ten genetic principal components of ancestry.

#### Non-linear analyses

Non-linear Mendelian randomization estimates were performed by first dividing the population into five equal-sized strata using the doubly-ranked method^25^. Estimates were then obtained in each stratum as per the linear analyses. Secondary analyses were performed by dividing the population into five equal-sized strata using the residual method for untransformed and log-transformed values of 25(OH)D concentrations^26^. The doubly-ranked method has been recommended for use with 25(OH)D concentrations, as in this case the genetic effect on the exposure is not constant in the population; this assumption is made by the residual stratification method^27^.

All statistical analyses were performed in R version 4.0.5.

### Ethics

UK Biobank ethical approval is provided by the UK Biobank research ethics committee and Human Tissue Authority research tissue bank. An independent Ethics and Governance Council oversees adherence to the Ethics and Governance Framework and provides advice on the interests of research participants and the general public in relation to UK Biobank. The current study was approved by UK Biobank (ref no. 98032).

## RESULTS

### Study population

A summary of participant characteristics is presented in Table 1. The mean age of participants was 57.1 years, and 53.4% of participants were female. Mean 25(OH)D concentration (mean-shifted to correspond to an October measurement) was 55.4 nmol/L. 2424 participants (0.7%) had fibromyalgia, 6764 (2.0%) had clinical fatigue, 4279 (1.3%) had chronic widespread pain, and 13,125 (3.9%) had probable lifetime major depression. The genetic instrument explained 4.7% of the variance in 25(OH)D levels, corresponding to an F statistic of 745.

**Table 1:** Participant characteristics for the analytic sample from UK Biobank

|  | UK Biobank |
| --- | --- |
| **Participants** | 333 025 |
| **Age at baseline** | 57.1 ± 8.1 |
| **Sex** |  |
| Female | 177 745 (53.4) |
| Male | 155 280 (46.6) |
| **25(OH)D concentration (nmol/L)** | 55.4 ± 19.3 |
| **Fibromyalgia cases** | 2424 (0.7) |
| **Clinical fatigue cases** | 6764 (2.0) |
| **Chronic widespread pain cases** | 4279 (1.3) |
| **Probable lifetime major depression cases** | 13 125 (3.9) |
| **BMI (kg/m<sup>2</sup>)** | 27.3 ± 4.8 |
| **SBP (mmHg)** | 137.5 ± 18.6 |
| **Smoking** |  |
| Current | 34 085 (10.2) |
| Other | 298 940 (89.8) |
| **Diabetes** |  |
| Known history | 15 822 (4.8) |
| No known history | 317 203 (95.2) |

Values represent mean ± standard deviation for continuous traits or N (%) for categorical traits. 25(OH)D concentrations are season-shifted to correspond to a measurement taken in autumn.

### Linear analyses

Linear Mendelian randomization estimates, representing the odds ratio for the average association with the outcome for 10 nmol/L higher genetically-predicted 25(OH)D in the whole population, are provided in Table 2. Estimates were 1.02 (95% confidence interval [CI]: 0.93, 1.12; p = 0.65) for fibromyalgia, 0.99 (95% CI: 0.94, 1.05; p = 0.86) for clinical fatigue, 0.95 (95% CI: 0.89, 1.02; p = 0.18) for chronic widespread pain, and 0.97 (95% CI: 0.93, 1.01; p = 0.18) for probable lifetime major depression.

**Table 2.** Linear Mendelian randomization estimates in the overall population

| Outcome | Estimate | 95% confidence interval | p-value |
| --- | --- | --- | --- |
| Fibromyalgia | 1.02 | 0.93, 1.12 | 0.65 |
| Clinical fatigue | 0.99 | 0.94, 1.05 | 0.86 |
| Chronic widespread pain | 0.95 | 0.89, 1.02 | 0.18 |
| Probable lifetime major depression | 0.97 | 0.93, 1.01 | 0.18 |

Estimates represent odds ratios per 10 nmol/L higher genetically-predicted concentration of 25(OH)D in the overall population.

### Non-linear analyses

Non-linear Mendelian randomization estimates, representing the odds ratio for the average association with the outcome for 10 nmol/L higher genetically-predicted 25(OH)D in each stratum of the population, are provided in Figure 1. As stratification is based on the exposure, the participants in each of the five strata (and hence the stratum-specific mean levels of 25(OH)D) are the same for each outcome. Associations were null in each stratum with the exception of chronic widespread pain in the stratum with the highest mean 25(OH)D levels (0.89, 95% CI: 0.80, 0.99; p=0.026), and probable lifetime major depression in the stratum with the lowest mean 25(OH)D levels (0.78, 95% CI: 0.64, 0.94; p = 0.009). A similar inverse association for probable lifetime major depression in this stratum was detected using the residual method for both untransformed and log-transformed 25(OH)D levels (Supplementary Table 2).

**Figure 1.** Non-linear Mendelian randomization estimates in strata of the population. Concentrations are stratum-specific mean levels of 25(OH)D. Estimates (95% confidence intervals) represent odds ratios per 10 nmol/L higher genetically-predicted concentration of 25(OH)D in strata of the population defined using the doubly-ranked method.

## DISCUSSION

We conducted one-sample Mendelian randomization analyses in UK Biobank using both linear and non-linear methods. We did not observe associations between genetically-predicted 25(OH)D and various outcomes in the population as a whole, suggesting that population shifts in vitamin D levels will not substantially decrease risk of fibromyalgia, clinical fatigue, chronic widespread pain, or probable lifetime major depression. However, we did observe an association with probable lifetime major depression in the stratum of the population with the lowest levels of 25(OH)D. This suggests that vitamin D supplementation may reduce risk of depression for those with low vitamin D levels.

Our findings may clarify evidence found in clinical trials for the effect of vitamin D supplementation on depression. While recent meta-analyses of randomized controlled trials of vitamin D supplementation have found evidence for reductions in depression and depressive symptoms^11-13^, the VITAL trial did not observe any reduction in risk^14^. However, few individuals in the VITAL trial had deficient levels of vitamin D. Additionally, VITAL trial participants were allowed to continue taking low dose vitamin D supplements, even if they were assigned to the placebo arm of the trial. Our findings suggest that any benefit of vitamin D supplementation may be limited to those with low vitamin D levels, and that supplementation beyond a threshold level may provide limited additional benefit. This corresponds to what has been seen in supplementation trials for acute respiratory tract infections, which have reported stronger evidence of risk reduction in those with 25(OH)D concentrations below 25 nmol/L^28^. Similarly, a previous systematic review of trials found that evidence of an inverse effect of vitamin D supplementation was limited to studies in those with deficient vitamin D levels at baseline^29^. This is also supported by research suggesting that vitamin D deficiency precedes the development of depression rather than being associated with its effects^30^.

We did not observe evidence suggesting any effect of vitamin D supplementation on fibromyalgia, clinical fatigue, or chronic widespread pain, beyond a slight protective association in one stratum that may represent a chance finding. This may be due to lack of power, as the number of cases for these outcomes was less than that for depression. Another challenge is defining these outcomes, as these conditions are not always accurately captured in routinely-collected healthcare records.

While our findings suggest that population-wide vitamin D supplementation will not substantially reduce risk of depression, they suggest vitamin D supplementation may reduce risk of depression for those with insufficient vitamin D levels. For any future clinical trials of vitamin D supplementation, this reinforces the importance of recruiting enough patients with vitamin D deficiency at baseline, so the trial has enough statistical power to detect an effect in deficient individuals^31^. However, recruiting enough vitamin D deficient individuals into such a trial to attain adequate power, and ensuring that those assigned to the placebo group do not self-medicate, may prove challenging in practice.

A number of different biological mechanisms might explain the direct relationship observed between low levels of circulating 25(OH)D and probable lifetime major depression. Inflammation is proposed to play a causal role in the development of depression^32, 33^, and vitamin D has been observed to play an anti-inflammatory role in neurological functioning by preventing oxidative damage to nervous tissue^34^. It has also been suggested in several randomized controlled trials that higher vitamin D levels result in lower levels of inflammatory cytokines^35^. There is also evidence that vitamin D plays a neuroprotective role in the proliferation, differentiation, survival and growth of neurons^36^. Recent studies in mice also indicate that a lack of vitamin D might influence microbiome diversity related to mood disorders^37^, in line with recent research into the role of gut microbiome in depression^38^. Finally, in vitro work has also suggested a connection between vitamin D and the hypothalamic–pituitary–adrenal (HPA) axis via influence on the glucocorticoid receptors^39^, which has a known role in the development of depression^40^. Given the number of biological pathways through which vitamin D levels might play a role in risk of depression, this may explain the associations we found between lower genetically-predicted 25(OH)D levels and probable lifetime major depression but not chronic pain conditions.

Our investigation has strengths in the Mendelian randomization design, large sample size, and biologically-driven choice of genetic variants. However, there are also potential limitations. Our analysis was restricted to individuals with European ancestry. This limits the validity and applicability of the findings, particularly as individuals with darker skin have lower 25(OH)D levels. Mendelian randomization analyses make the assumption that the only causal pathway from the genetic variants to the outcome is via the exposure. While the four gene regions chosen for this study have strong biological links to vitamin D, this assumption cannot be tested empirically. Further, even if the Mendelian randomization assumptions are satisfied, genetic variants may influence 25(OH)D concentrations in a different way to dietary supplementation or other clinical interventions. We used the doubly-ranked method for non-linear Mendelian randomization, which is more robust than the residual method, but some residual bias owing to variability in the genetic effects on the exposure may remain^27^. Our outcome definitions are likely to have missed some cases; we only identified around 4% of UK Biobank participants as having depression, whereas lifetime prevalence of major depression is around 10-20% according to general population studies^41^. Misclassification of cases as controls generally introduces a bias towards the null, so the genetic associations observed in our analysis are likely to be conservative, and hence may underestimate the true impact on disease risk.

In conclusion, we found evidence for an effect of vitamin D on depression in individuals with low vitamin D status, but no evidence of an effect at any level of 25(OH)D for other pain outcomes.

## Data Availability

UK Biobank data are available on reasonable request to any bona fide researcher.

## ACKNOWLEDGEMENTS

SSZ is supported by a National Institute for Health Research Clinical Lectureship and works in centres supported by Versus Arthritis (grant no. 21173, 21754 and 21755). SB is supported by the Wellcome Trust (225790/Z/22/Z) and the United Kingdom Research and Innovation Medical Research Council (MC_UU_00002/7). This research was supported by the National Institute for Health Research Cambridge Biomedical Research Centre (NIHR203312). The views expressed are those of the authors and not necessarily those of the National Institute for Health Research or the Department of Health and Social Care.

## CONFLICTS OF INTEREST

The authors declare no conflict of interest.

## SUPPLEMENTARY INFORMATION

Supplementary information is available at Molecular Psychiatry’s website

## Notes

### Competing Interest Statement

The authors have declared no competing interest.

