## Supplemental table 1 for "Vitamin D, chronic pain, and depression: linear and non-linear Mendelian randomization analyses"

**Supplementary Table 1:** List of genetic variants for allelic score

| Chromosome: Position (hg19) | rsID | Effect allele | Other allele | Conditional association with 25(OH)D (nmol/L) |
| --- | --- | --- | --- | --- |
| 4:72617775 | rs1352846 | G | A | 0.172 |
| 4:72618334 | rs7041 | C | A | -0.045 |
| 4:72634343 | rs4694431 | T | C | -0.034 |
| 4:72770563 | rs139148694 | GTGCTTTTATCAA | G | 0.028 |
| 11:14339328 | rs16913816 | A | G | -0.031 |
| 11:14900931 | rs117913124 | A | G | 0.503 |
| 11:14912573 | rs117576073 | T | G | 0.246 |
| 11:14913575 | rs12794714 | A | G | 0.139 |
| 11:14913645 | rs202122669 | A | G | -0.615 |
| 11:14913900 | rs187639972 | C | G | -0.360 |
| 11:14941652 | rs117115472 | G | C | 0.148 |
| 11:71157867 | rs139168803 | A | G | -0.188 |
| 11:71158672 | rs12573951 | G | A | -0.045 |
| 11:71161063 | rs7928249 | G | A | -0.131 |
| 11:71180762 | rs549000212 | A | C | -0.364 |
| 11:71290740 | rs4081429 | C | A | 0.017 |
| 20:52714706 | rs6123359 | G | A | -0.026 |
| 20:52731402 | rs6127099 | T | A | 0.013 |
| 20:52735238 | rs35870583 | GT | G | 0.027 |
| 20:52737123 | rs2585442 | G | C | -0.025 |
| 20:52788925 | rs2762942 | A | G | -0.053 |
