## Supplemental table 2 for "Vitamin D, chronic pain, and depression: linear and non-linear Mendelian randomization analyses"

**Supplementary Table 2:** Non-linear Mendelian randomization estimates for probable lifetime major depression from different methods.

|  | Doubly-ranked method | | Residual method (untransformed exposure) | | Residual method (log-transformed exposure) | |
| --- | --- | --- | --- | --- | --- | --- |
| Stratum | Estimate | 95% CI | Estimate | 95% CI | Estimate | 95% CI |
| 1 (lowest) | 0.78 | 0.64, 0.94 | 0.86 | 0.75, 0.99 | 0.91 | 0.82, 1.01 |
| 2 | 0.98 | 0.86, 1.12 | 0.94 | 0.85, 1.03 | 0.94 | 0.85, 1.03 |
| 3 | 0.95 | 0.86, 1.05 | 0.97 | 0.88, 1.06 | 1.02 | 0.92, 1.12 |
| 4 | 1.05 | 0.98, 1.14 | 1.00 | 0.92, 1.10 | 0.97 | 0.88, 1.07 |
| 5 (highest) | 0.98 | 0.92, 1.05 | 1.01 | 0.93, 1.09 | 1.02 | 0.92, 1.12 |

Estimates (95% confidence intervals) represent odds ratios per 10 nmol/L higher genetically-predicted concentration of 25(OH)D (for doubly-ranked method and residual method with untransformed exposure), or per 20% higher genetically-predicted concentration of 25(OH)D (for residual method with log-transformed exposure).
